# Lifetime risk of maternal near miss morbidity: A novel indicator of maternal health

**DOI:** 10.1101/2023.03.09.23287053

**Authors:** Ursula Gazeley, Julio Romero Prieto, José Manuel Aburto, Antonino Polizzi, Georges Reniers, Veronique Filippi

## Abstract

The lifetime risk of maternal death is the risk that a 15-year-old girl will die of a maternal cause in her reproductive lifetime. Its intuitive appeal means it is a widely used summary measure for advocacy and international comparisons of maternal health. But relative to mortality, women are at a higher risk of experiencing life-threatening maternal morbidity called “maternal near miss” events – complications so severe that women almost died. As maternal mortality continues to decline, stronger health indicators that include information on both fatal and non-fatal maternal outcomes are required. Thus, we propose a novel measure – the lifetime risk of maternal near miss – to estimate the risk a girl will experience at least one maternal near miss in her reproductive lifetime, accounting for survival from ages 15-49. This new indicator is urgently needed because existing measures of maternal morbidity prevalence (near miss ratio or rate) do not capture the cumulative risk over the reproductive life course. We use estimates of fertility and survival from the World Population Prospects for Kenya in 2021 along with simulated data on the maternal near miss ratio to demonstrate the calculation of the lifetime risk of maternal near miss. We estimate that the lifetime risk of maternal near miss in Kenya is 1 in 37, compared to a lifetime risk of maternal death of 1 in 59.

**Key messages:**

- We propose a new indicator – the lifetime risk of maternal near miss – to estimate the risk of a 15-year-old girl experiencing a severe life-threatening maternal complication over her reproductive life course, accounting for survival between the ages 15-49.
- This indicator is needed because no existing measure of maternal near miss morbidity prevalence (ratio or rate) accounts for the cumulative risk of severe complications with each pregnancy.
- We demonstrate two methods for the calculation of the lifetime risk of maternal near miss, the choice of which depends on whether (i) estimates of the maternal near miss ratio by age group or, (ii) a summary estimate for ages 15-49 years, are available.
- We advocate for the use of this indicator to compare trends in maternal near miss morbidity alongside trends in maternal mortality.

## Introduction

The lifetime risk of maternal death (LTR-MD) is a widely used summary measure of maternal health. As most commonly measured, this denotes the probability of a 15-year-old girl dying from a maternal cause in her remaining reproductive lifetime, accounting for other causes of mortality (1). Its intuitive appeal means it is used to compare countries and changes over time in World Health Organization (WHO) and United Nations joint maternal mortality estimates (2). However, maternal deaths are just the tip of the iceberg of poor maternal outcomes. For every woman that dies from a maternal cause, as many as 20 women may experience a life-threatening maternal near miss complication (3), with potential implications for women’s long-term survival and physical and mental health outcomes (3–8). The WHO define a maternal near miss as a *“woman who nearly died but survived a complication that occurred during pregnancy, childbirth, or within 42 days of termination of pregnancy”* (9). Cases are identified on the basis of the clinical, laboratory, and management-based criteria of organ dysfunction (9).

We propose a new indicator – the lifetime risk of maternal near miss (LTR-MNM) – to measure the risk of experiencing a maternal near miss during a woman’s reproductive lifetime. This novel metric is required because existing measures of the frequency of maternal near miss in relation to either the number of live births (MNMRatio) or to the female population of reproductive age (MNMRate) do not quantify the cumulative risk over women’s lives, which is dependent on the number of times she is exposed (i.e., fertility levels). Nor do they capture how the risk of experiencing a maternal near miss during the reproductive life course is dependent upon a woman surviving her reproductive years. We need a new summary measure to go beyond estimating the risk of each pregnancy and instead capture the impact of maternal near miss morbidity across the female reproductive life course. As a function of the maternal near miss ratio, fertility, and mortality levels, the LTR-MNM addresses this deficit and captures these potentially countervailing dynamics.

As countries advance through the obstetric transition, fewer women die from maternal causes (10). Expansion in access to and improvements in the quality of emergency obstetric care means that many more women who would have otherwise died in the community now survive pregnancy and the immediate 42 day postpartum period (10). This change has motivated international-level calls for a stronger indicator of maternal morbidity for monitoring purposes.

Using the equation for the LTR-MD as a starting point, we present a formula for the calculation of the LTR-MNM. We then describe the step-by-step calculation of the LTR-MNM for Kenya – a country with a high burden of maternal mortality and morbidity (2) – combining simulated data on the maternal near miss ratio, and fertility and survival data from the World Population Prospects for Kenya 2021. Finally, we discuss the strengths and limitations of our proposed indicator.

## Basic concepts

### 1. Maternal mortality and the LTR-MD

As described by Wilmoth et al (1), the lifetime risk of maternal death (LTR-MD) can be calculated using the Maternal Mortality Ratio (MMRatio: the number of maternal deaths at age *x* per 1000 live births at maternal age *x*) or the Maternal Mortality Rate (MMRate: the number of maternal deaths per 1000 woman-years lived at maternal age x) as follows:

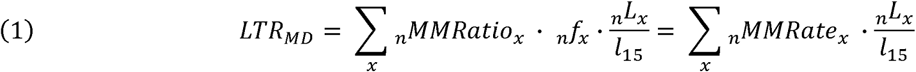

where _*n*_*f*_*x*_ is the fertility rate between ages *x* and *x+ n* (where n is the length of the age interval), 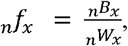, where _*n*_*B*_*x*_ is the number of live births for women aged *x* to *x+ n*, and _*n*_*W*_*x*_ is the number of woman-years of exposure for ages *x* to *x+ n*, in the observed population; _*n*_*L*_*x*_ is the number of woman-years of exposure to the risk of dying from maternal or other causes between ages *x* and *x+ n*, and *l*_15_ is the probability that a girl will survive to age 15. Both _*n*_*L*_*x*_ and *l*_15_ are life table measures. Equation (2) is summed across all reproductive ages, (e.g. 15 to 49 years) to calculate the cumulative risk of maternal death across the female reproductive life course.

Using period data, the LTR-MD quantifies the risk of death from a maternal cause in a synthetic cohort, conditional on survival to age 15, with other competing causes of mortality considered. Further detail on the concept and measurement of the lifetime risk of maternal death can be found in Wilmoth et al (2009) (1).

### 2. Maternal morbidity and the lifetime risk of maternal near miss morbidity

We propose that there is utility in extending the concept of lifetime risk to measure the cumulative risk of maternal near miss morbidity (LTR-MNM) throughout the reproductive life course. As is the case with the LTR-MD (1), the LTR-MNM can be calculated using either the (i) the maternal near miss ratio (MNMRatio: the number of maternal near miss per 1000 live births) or (ii) the maternal near miss rate (MNMRate: the number of maternal near miss per 1000 woman-years lived). As the MNMRatio is more frequently available, we use this to calculate the LTR-MNM as follows:

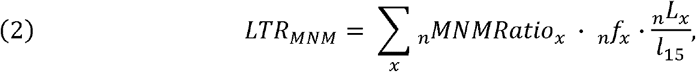

Equation 2 gives a hypothetical measure of the risk of experiencing a maternal near miss during the reproductive life course, conditional on survival to age 15, and accounting for survival from age 15-49. This is the risk of experiencing at least one maternal near miss, though in reality a woman may experience more than one near miss in her reproductive lifetime. As women with a maternal near miss would have died without receiving care at the facility, the numerator of the MNMRatio can be hypothesised as representing the true number of cases in a community. However, to avoid overestimating the MNMRatio and hence the LTR-MNM, the denominator should estimate live births in the community and not only in the facility – especially when the prevalence of institutional delivery is low.

Below we describe the calculation of the LTR-MNM depending on the availability (or not) of age-disaggregated estimates of the MNMRatio. All procedures were conducted using the R software (11) and are fully reproducible from open data. Our code is posted in a public code repository: https://osf.io/8brgp/?view_only=19cea2858df94b2b877d4ff57cfa7e48.

#### Calculation when (abridged) age-specific maternal near miss data are available

The following example for the calculation of the LTR-MNM in Kenya in 2021 uses simulated data for the MNMRatio. For all calculations, we assumed an overall MNMRatio for ages 15-49 combined of 8.38 per 1000 live births. This is the average of four recent studies of the prevalence of MNM in Kenya (12–15). We used data from Kenya on total births by five-year age groups from the World Population Prospects 2021, adjusted for a stillbirth rate of 19 per 1000 (16), to simulate possible age distributions of maternal near miss cases and MNMRatio because age-disaggregated data were not available in Kenya. Following evidence on the MNMRatio by age group from Brazil (17), and global evidence on the risk of maternal death by age (15), we hypothesised that a “j-shaped” risk profile might be most plausible and was used for the worked example: a slightly higher risk for adolescent ages 15-19, falling to a minimum at ages 20-24, and increasing with increasing maternal age thereafter.

The MNMRatio for single year-age groups are virtually never available, and hence are not described here. Calculation of the LTR-MNM by five-year age groups assumes the MNMRatio (and fertility and survival) is constant throughout each age interval – a simplifying assumption, but one that better reflects the best-case scenario for real-world near miss data. Estimates of age-specific fertility rates, _*n*_*f*_*x*_, survivors to age 15, *l*_15_, and the number of woman-years lived in the interval, _*n*_*L*_*x*_, by five-year age group derive from the World Population Prospects abridged life tables for Kenya in 2021 (18). Data are open access and can be downloaded from the World Population Prospects website. Table 1 presents the simulated MNMRatio data, the WPP fertility and survival data, and the calculation of the LTR-MNM by each five-year age group.

**Table 1.**
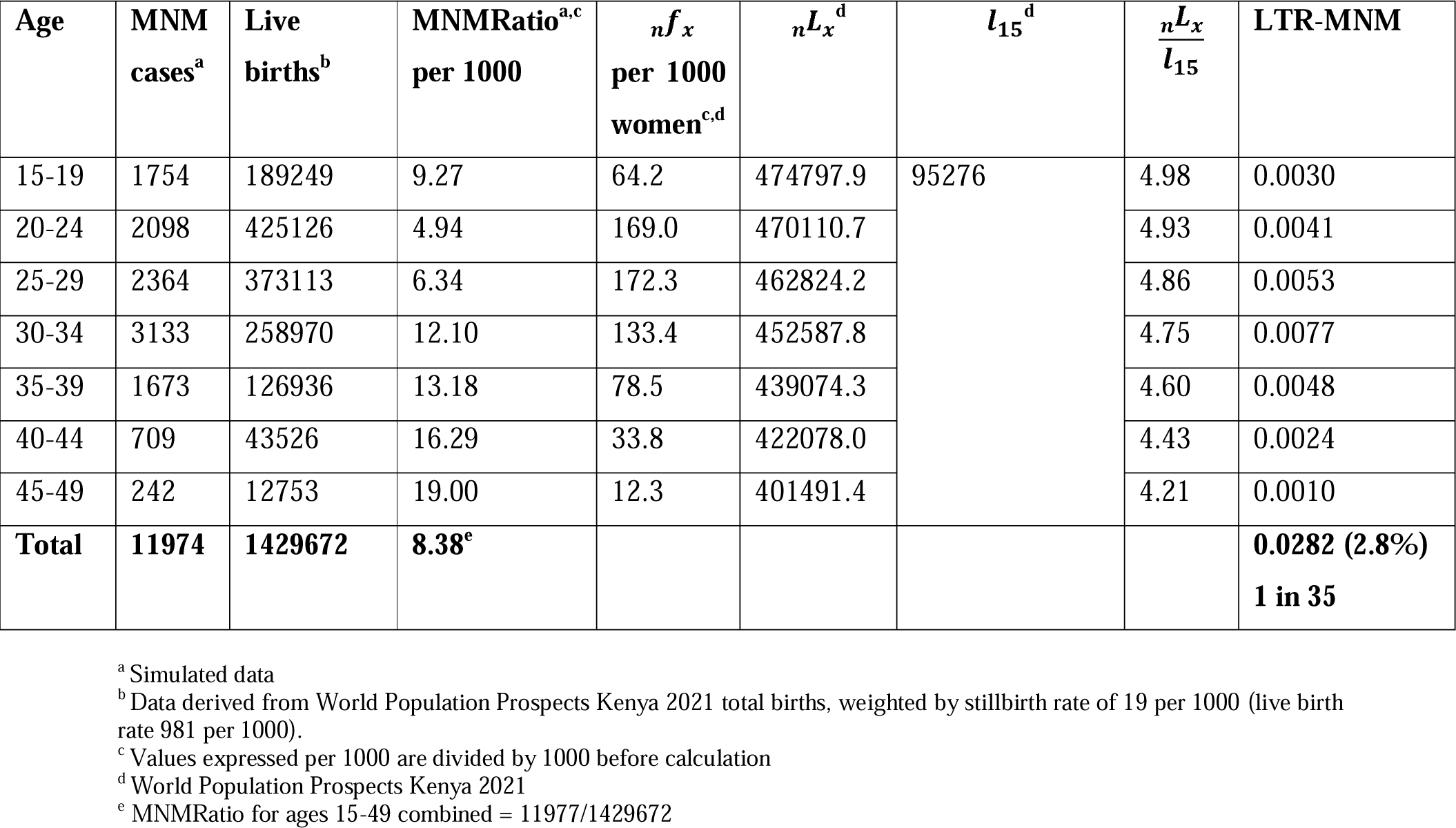
LTR-MNM Kenya 2021 calculation assuming “j-shaped” MNM age distribution.

| Age | MNM cases<sup>a</sup> | Live births<sup>b</sup> | MNMRatio<sup>a,c</sup> per 1000 | ${}_nf_x$ per 1000 women<sup>c,d</sup> | ${}_nL_x^d$ | $l_{15}^d$ | $\frac{{}_nL_x}{l_{15}}$ | LTR-MNM |
| --- | --- | --- | --- | --- | --- | --- | --- | --- |
| 15-19 | 1754 | 189249 | 9.27 | 64.2 | 474797.9 | 95276 | 4.98 | 0.0030 |
| 20-24 | 2098 | 425126 | 4.94 | 169.0 | 470110.7 |  | 4.93 | 0.0041 |
| 25-29 | 2364 | 373113 | 6.34 | 172.3 | 462824.2 |  | 4.86 | 0.0053 |
| 30-34 | 3133 | 258970 | 12.10 | 133.4 | 452587.8 |  | 4.75 | 0.0077 |
| 35-39 | 1673 | 126936 | 13.18 | 78.5 | 439074.3 |  | 4.60 | 0.0048 |
| 40-44 | 709 | 43526 | 16.29 | 33.8 | 422078.0 |  | 4.43 | 0.0024 |
| 45-49 | 242 | 12753 | 19.00 | 12.3 | 401491.4 |  | 4.21 | 0.0010 |
| **Total** | **11974** | **1429672** | **8.38<sup>e</sup>** |  |  |  |  | **0.0282 (2.8%)**<br>**1 in 35** |

<sup>a</sup> Simulated data
<sup>b</sup> Data derived from World Population Prospects Kenya 2021 total births, weighted by stillbirth rate of 19 per 1000 (live birth rate 981 per 1000).
<sup>c</sup> Values expressed per 1000 are divided by 1000 before calculation
<sup>d</sup> World Population Prospects Kenya 2021
<sup>e</sup> MNMRatio for ages 15-49 combined = 11977/1429672

The steps followed in this calculation are as follows:

1. For each age group, the MNMRatio is multiplied by the age-specific fertility rates, _*n*_*f*_*x*_ (this gives the MNMRate).
2. This is then multiplied by the survival fraction 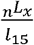 – person-years lived throughout the interval divided by the number of female survivors to age 15.
3. The resulting estimates of the LTR-MNM for each five-year age group are then summed to get the final LTR-MNM.
4. Finding the reciprocal of this total allows us to express the LTR-MNM as a 1 in n risk.

In this simulated example, the resulting LTR-MNM is 1 in 35, such that conditional on surviving to age 15, a girl would face a 1 in 35 chance of experiencing a maternal near miss complication during her reproductive lifetime, conditional on survival from ages 15-49. This compares to a LTR-MD of 1 in 59 (see Lifetime Risk of Severe Maternal Outcome, below).

#### Sensitivity of the LTR-MNM estimate to the age-distribution of the MNMRatio

The worked example above, we assumed a “j-shaped” age profile for the MNMRatio. In reality, even at a fixed level of maternal morbidity for reproductive ages 15-49 combined (8.38 per 1000 live births), the age pattern of the MNMRatio could adopt a variety of shapes (e.g. u-shaped, n-shaped, constant, decreasing, and increasing). Figure 1 shows simulated MNM age distributions, and Table 2 shows the resulting estimates of the LTR-MNM. Despite substantial differences in the underlying MNMRatio by age group, the resulting LTR-MNM for all ages combined are similar. Full calculations of the LTR-MNM for each age distribution can be found in the Supplementary Material (Table S1).

**Table 2.** Sensitivity of LTR-MNM for Kenya 2021 to simulated MNM age distribution.

| Age distribution of MNM | LTR-MNM | LTR-MNM 1 in n |
| --- | --- | --- |
| J shape | 0.0282 | 1 in 35 |
| N shape | 0.0282 | 1 in 35 |
| U shape | 0.0260 | 1 in 38 |
| Constant | 0.0267 | 1 in 37 |
| Decreasing | 0.0256 | 1 in 39 |
| Increasing | 0.0284 | 1 in 35 |

**Figure 1.**
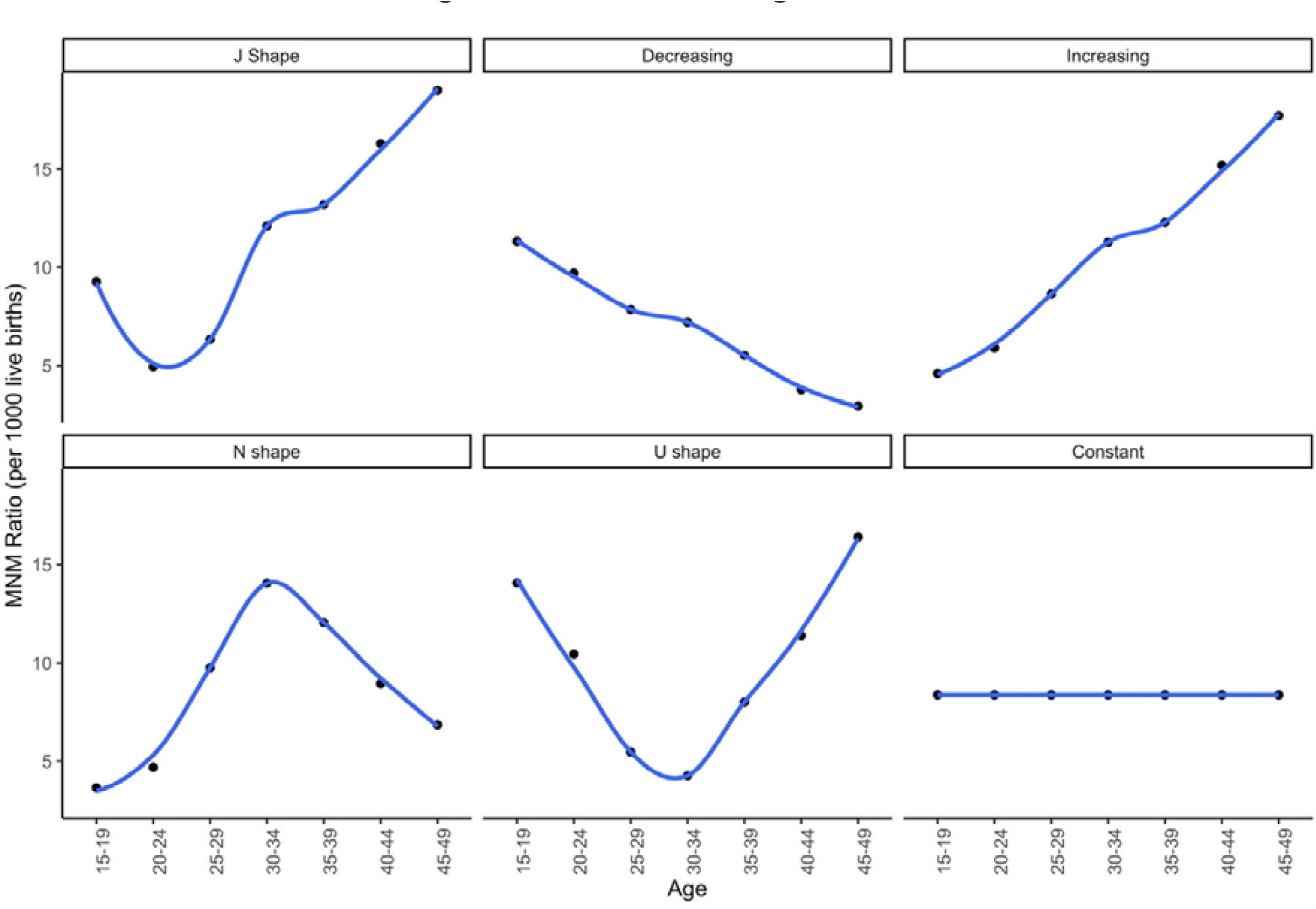
Simulated MNM age distributions. Note: all distributions have a MNMRatio for ages 15-49 combined of 8.38 per 1000 live birth

#### Calculation when only a summary estimate of maternal near miss for all ages 15-49 is available

Age-specific maternal near miss data are often not available. Rather, an estimate of the MNMRatio is often calculated for all reproductive ages combined. When this is the case, the LTR-MNM can still be calculated, though this relies on the assumption that the risk of MNM is constant throughout the reproductive ages. This is a simplifying assumption most appropriate for data-scarce contexts, or when data processing has resulted in a loss of detail. As in the case of LTR-MD, if we assume the MNMRatio is constant throughout the reproductive ages, equation (2) becomes:

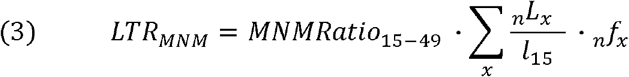

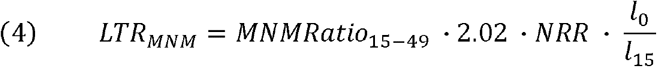

Where *l*_0_ is the initial female population radix (100,000) and the NRR is the net reproduction rate (the expected number of female children per newborn girl). This is scaled by a factor of 2.02, assuming a sex ratio at birth of 102 boys to 100 girls in Kenya (18), because the NRR is expressed in terms of female births only, not total births, _*n*_*f*_*x*_. Assuming an NRR of 1.5114 (WPP Kenya 2021) (18), applying formula 4, the LTR-MNM is:

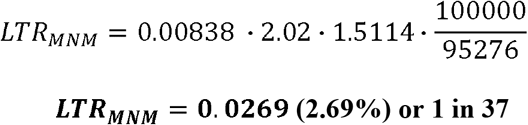

This summary estimate of the LTR-MNM for ages 15-49 combined falls in the middle of the results for different possible age distributions above (1 in 39 to 1 in 35) and suggests that equation (4) is a reasonable approximation where age-disaggregated maternal near miss data are not available.

#### Lifetime risk of severe maternal outcome

The concept of LTR-MNM can be used in addition to the LTR-MD to estimate the lifetime risk of severe maternal outcome (LTR-SMO). As SMO is the summation of MNM and maternal deaths, the LTR-SMO becomes:

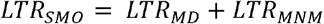

Assuming an MMRatio of 530 per 100,000 for ages 15-49 in Kenya (2), corresponding to a LTR-MD of 0.017 or 1 in 59 (using equation 4 with the MMRatio), the LTR-SMO becomes:

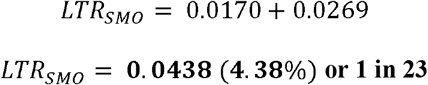

This means there is a 1 in 23 risk of a 15-year-old girl experiencing either a maternal death or a life-threatening MNM complication during her reproductive lifetime. Maternal near miss morbidity accounts for 61% of the LTR-SMO in this example (LTR-MNM/(LTR-MD + LTR-MNM)). This relative contribution will vary depending on a country’s position in the obstetric transition.

#### Uncertainty

Where estimates of the number of maternal near miss cases derive from sample surveys, estimates of the MNMRatio and hence the LTR-MNM are subject to sampling variability. In frequentist models, the 95% confidence interval for the MNMRatio could be used to calculate corresponding uncertainty in the LTR-MNM. In Bayesian models, such as joint WHO and UN agency reports on the trends in maternal mortality, an 80% uncertainty interval for the MMRatio and LTR-MD is estimated using the 10^th^ and 90^th^ percentile of the posterior distribution (19). Either approach could be replicated for the LTR-MNM.

### Strengths and limitations

#### Strengths

The LTR-MNM is a novel concept to estimate the risk that a 15-year-old girl will experience at least one maternal near miss complication in her reproductive lifetime. Unlike existing measures of near miss prevalence (e.g., ratio or rate), this indicator quantifies the cumulative risk of experiencing maternal near miss morbidity across the female reproductive life course. Maternal near miss complications are a key adverse pregnancy outcome, with potentially significant effects on women’s future mental and physical health outcomes. As the obstetric transition progresses and fewer women die of maternal causes, the LTR-MNM is essential to strengthen international monitoring of maternal morbidity and advocacy for quality emergency obstetric care.

Second, differences in the LTR-MNM between populations and over time can also be decomposed into changes in the risk of near miss with each pregnancy (MNMRatio), the number of times women are exposed (_*n*_*f*_*x*_), and changes to all-cause mortality (e.g., including mortality shocks that affect the _*n*_*L*_*x*_ schedule such as COVID-19). Disentangling these dynamics can improve our understanding of the global burden of maternal morbidity across the female reproductive life course.

Though availability of data to estimate the MNMRatio is often poor, especially in low resource settings where the burden of maternal morbidity is highest, we have shown that the summary-level estimate of the LTR-MNM falls within the range of estimates derived from age-specific data. If the risk of near miss increases after a certain age as is the case with the risk of maternal death (20) (i.e., according to a j-shape, u-shape or increasing pattern in Figure 1) the summary estimate of LTR-MNM may be an underestimate and is therefore best interpreted as a lower bound on the true cumulative risk of maternal near miss morbidity.

### Limitations

In analogy with the (period) life expectancy and the LTR-MD, the LTR-MNM is a synthetic cohort measure of population health and presumes that women experience today’s maternal morbidity, mortality, and fertility conditions throughout their lifetime i.e., rates observed in a particular year are assumed to be constant for future cohorts. For this reason, it cannot be interpreted as a prediction of the lifetime risk of a maternal near miss in a real cohort because the maternal near miss risk may change (decline) in the future. Its dependence on life table measures does, however, mean it is a comparable indicator over time and across populations. Second, women who experience a maternal near miss may face elevated mortality risks and therefore have a lower _*n*_*l*_*x*_ schedule. This would cause us to overestimate the LTR-MNM. Third, experiencing a near miss is a potentially repeating, non-independent event because having an initial near miss may increase a woman’s future risk of experiencing a subsequent near miss. This calculation does not account for potential heterogeneity but amounts to a population average of the risk of experiencing at least one maternal near miss during the reproductive life course. Finally, estimates of the MNMRatio often derive from primarily one-off facility-based surveys which makes national-level estimation of the LTR-MNM difficult. Further work is required to inform the aggregation of maternal near miss estimates (and MNMRatio) to produce nationally representative estimates of the LTR-MNM.

## Conclusion

We propose the lifetime risk of maternal near miss as a much-needed new summary measure of maternal health, in addition to mortality. Comparability of estimates would benefit from improvements in the national-level aggregation of maternal near miss, especially in high burden settings.

## Data Availability

All data used in this article are freely available for download from the UN World Population Prospects Download Center: https://population.un.org/wpp/Download/Standard/MostUsed/. All code used in this analysis is available from the Open Science Framework repository: https://osf.io/8brgp/?view_only=19cea2858df94b2b877d4ff57cfa7e48 .

## Declarations

### Ethical approval

Ethical approval was not required as to demonstrate our new indicator we used population-level data available in the public domain (World Population Prospects).

### Data availability

All data used in this article are freely available for download from the UN World Population Prospects Download Center: https://population.un.org/wpp/Download/Standard/MostUsed/. All code is available from the Open Science Framework repository: https://osf.io/8brgp/?view_only=19cea2858df94b2b877d4ff57cfa7e48.

### Supplementary data

Supplementary data are available at IJE online.

### Author contributions

UG conceived the idea, performed the computations, and prepared the results. JRP, JMA, AP, GR and VF supported the interpretation of results and the refinement of the simulations. AP developed the code repository and refined the code. UG drafted the initial manuscript. JRP, JMA, AP, GR and VF revised the article.

