## Supplementary Table 1 for "Lifetime risk of maternal near miss morbidity: A novel indicator of maternal health"

**Contents:** Table S1 provides the full lifetime risk of maternal near miss (LTR-MNM) calculation for each simulated age-profile of maternal near miss (MNM) risk for Kenya.

**Table S1: LTR-MNM Kenya 2021 calculation for each simulated MNM age distribution**

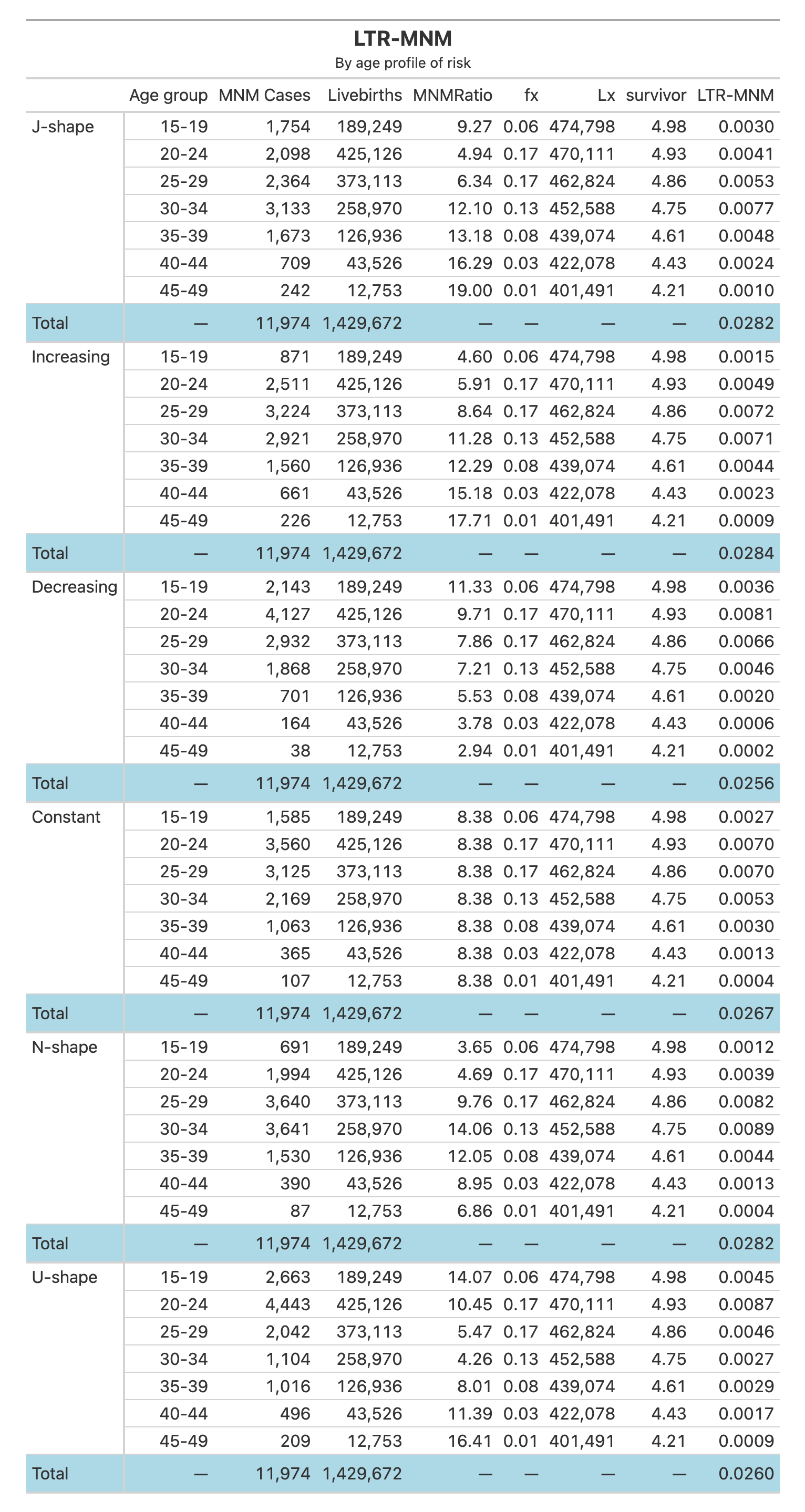
